# Automatic detection of pituitary microadenoma from magnetic resonance imaging using deep learning algorithms

**DOI:** 10.1101/2021.03.02.21252010

**Authors:** Qingling Li, Yanhua Zhu, Minglin Chen, Ruomi Guo, Qingyong Hu, Zhenghui Deng, Songqing Deng, Huiquan Wen, Rong Gao, Yuanpeng Nie, Haicheng Li, Tiecheng Zhang, Jianning Chen, Guojun Shi, Jun Shen, Wai Wilson Cheung, Yulan Guo, Yanming Chen

**Author notes:** Corresponding author: Yanming Chen, Department of Endocrinology and Metabolism, The Third Affiliated Hospital, Sun Yat-Sen University, 600 Tianhe Road, Guangzhou, China, 510630., Yulan Guo, School of Eletronics and Communication Engineering, Sun Yat-Sen University, 135 Newport West Road, Guangzhou, China, 510275. These authors contributed equally to this work (Qingling Li, Yanhua Zhu, Minglin Chen, Ruomi Guo). **Author contributions:** Guarantors of integrity of entire study, Y.M.C., Y.L.G., study concepts/study design or data acquisition or data analysis/interpretation, all authors, manuscript drafting or manuscript revision for important intellectual content, all authors, approval of final version of submitted manuscript, all authors.

## Abstract

Pituitary microadenoma (PM) is often difficult to detect by MR imaging alone. We employed a computer-aided PM diagnosis (PM-CAD) system based on deep learning to assist radiologists in clinical workflow. We enrolled 1,228 participants and stratified into 3 non-overlapping cohorts for training, validation and testing purposes. Our PM-CAD system outperformed 6 existing established convolutional neural network models for detection of PM. In test dataset, diagnostic accuracy of PM-CAD system was comparable to radiologists with > 10 years of professional expertise (94% versus 95%). The diagnostic accuracy in internal and external dataset was 94% and 90%, respectively. Importantly, PM-CAD system detected the presence of PM that had been previously misdiagnosed by radiologists. This is the first report showing that PM-CAD system is a viable tool for detecting PM. Our results suggest that PM-CAD system is applicable to radiology departments, especially in primary health care institutions.

## INTRODUCTION

Prevalence of pituitary adenomas (PA) ranges from 1 in 865 to 1 in 2688 adults. PA may hypersecrete hormones or cause mass effects, which result in various clinical symptoms, including infertility, diabetes insipidus and hypopituitarism ^[1]^. The end of the one-child policy and social shifts in China have resulted in the increasing demand for fertility treatments. Pituitary tumors, although usually benign, can inhibit the production of follicle-stimulating hormone (FSH) or luteinizing hormone (LH) and cause infertility. Functional PA have been found in many infertile patients, the most common is prolactinoma. Approximately 50% of all pituitary adenomas and 90% of prolactinomas are microadenomas, that rarely increase in size^[2,3]^. Timely diagnosis and follow-up of pituitary microadenoma (PM), especially functional PM, is particularly important ^[4]^. Due to its relatively small in size and variable anatomical structure among individuals, the diagnosis of PM is not easy by applying the technique of magnetic resonance imaging (MRI) alone ^[1,5]^. Manual analysis of MRI data is usually difficult, biased and time-consuming, and the diagnostic accuracy is closely related to the radiologist’s experience. Therefore, MRI-based diagnosis for PM needs to be improved ^[6,7]^. The increase in the demand for radiologist is an inevitable trend, but the supply of radiologist has not increased proportionally ^[8]^. A shortage of radiologists restricts the continuity in radiology services and causes a delay in diagnosis, compromising the overall quality of service to patients ^[8, 9]^. Recently, we have encountered several cases of misdiagnosed PM in our hospital (Fig 1). Deep learning has dominated various computer vision areas since 2012 due to its effective representation capability ^[10]^. The development of convolutional neural network (CNN) has significantly improved the performance of image classification and object detection ^[11]^. Deep learning has the potential to revolutionize disease diagnosis and management by improving the diagnostic accuracy and reducing the workload of clinicians. Specifically, CNN has achieved great progresses in the diagnosis of breast cancer ^[12,13]^, diabetes retinopathy ^[14,15]^, fibrotic lung disease ^[16]^ and COVID-19 ^[17]^. Furthermore, reports demonstrated that computer-aided diagnosis (CAD) system can accurately diagnose patients with PA from MR images ^[18-20]^. However, the information for CAD-based diagnosis of PM is limited ^[5,6]^. In this work, we have constructed a computer-aided PM diagnosis (PM-CAD) system based on deep learning and aimed to provide an accurate and timely diagnosis of PM from pituitary MR images.

**Fig 1.**
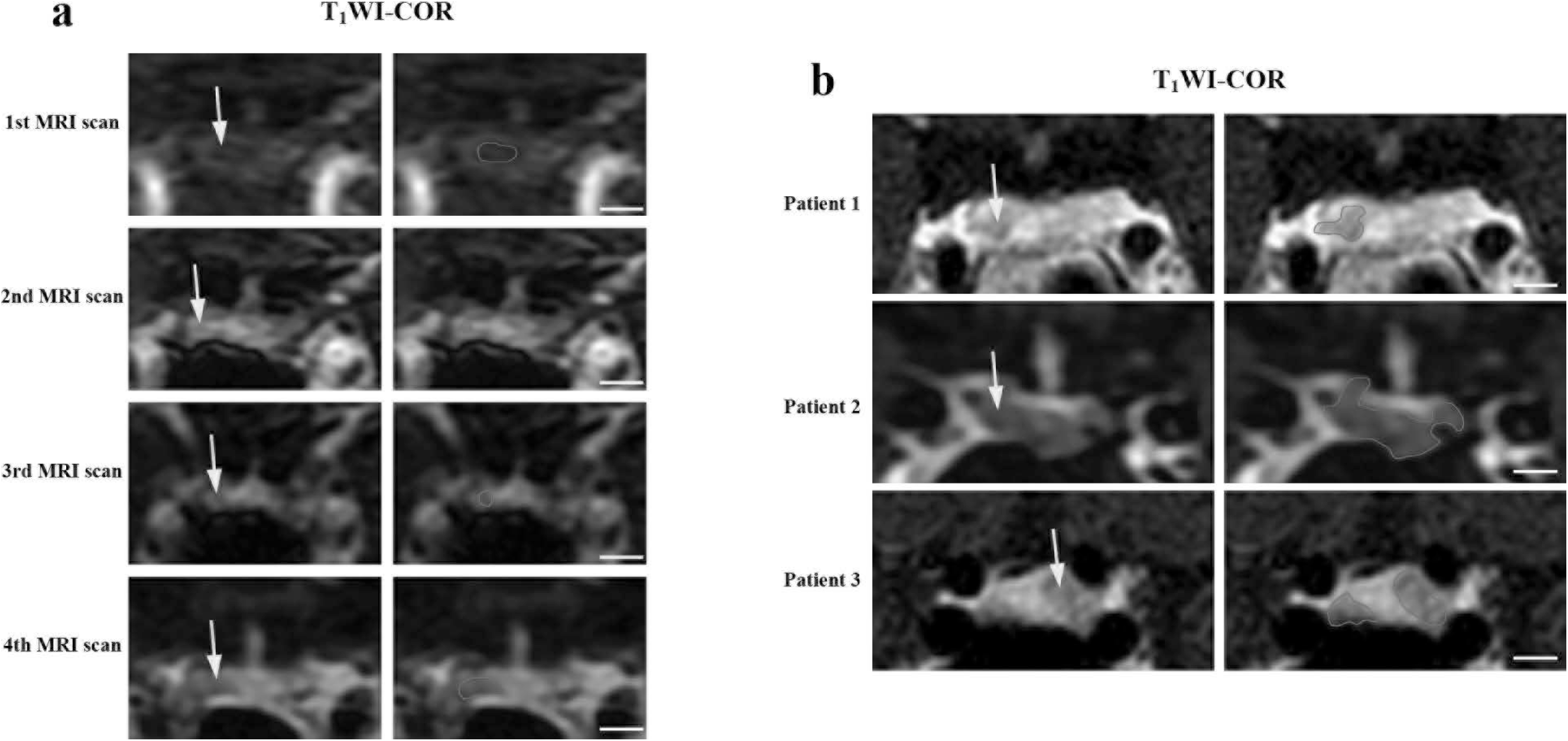
Cases of 4 misdiagnosed pituitary microadenoma. (a) 4 consecutive pituitary MRI scans over a period of 20 months in a misdiagnosed patient with pituitary microadenoma. The radiologists have not detected the pituitary microadenoma during the first 3 MRI examinations. A functional microadenoma has been localized by the subsequent ACTH examination of the inferior petrosal sinus in the region of right pituitary gland. On the 4th MRI scanning, two microadenoma are detected by radiologist. (b) Additional 3 cases of misdiagnosed microadenoma. Patient 1 has a very small microadenoma with a diameter < 3 mm. Patient 2 has an irregularly shaped microadenoma. Patient 3 has two microadenoma (with diameters of 2.8 mm and 6.1 mm, respectively) and the smaller one was misdiagnosed. The comprehensive clinical data for patients were listed in Supplementary Table 1. ACTH: Adrenocorticotropic Hormone. MRI: magnetic resonance imaging. T1WI-COR: T1 weighted imaging-coronal. MRI bar = 5 mm. The yellow arrow and the area inside the red circle represent adenomas.

## RESULTS

### Cases of misdiagnosed pituitary microadenoma in our hospital

Our current research in applying deep learning algorithms to aid in the diagnosis of PM was prompted by several misdiagnosed cases in our hospital. A female patient in her 40’s suffered from diabetes, osteoporosis and recurrent cellulitis. Laboratory examination and functional test were consistent with the presentation of Cushing’s disease (Tab S1). The patient underwent three times of MRI scanning over a period of 20 months. However, the radiologists did not detect any pituitary adenoma (Fig 1a). Nevertheless, this patient was given bromocriptine and cyproheptadine for 2 years, but the magnitude of the disease was not under effective control. Subsequently, the inferior petrosal sinus sampling (IPSS) was performed on this patients and a functional microadenoma was finally detected in her right pituitary gland. We performed another MRI examination on this patient and were able to detect 2 pituitary microadenomas (with diameters of 5 mm and 3 mm, respectively) (Fig 1a). One week later, we performed transsphenoidal PM resectioning on this patient. Shortly after the completion of the operation, the serum concentration of ACTH and free cortisol of this patient was normalized (Tab S1). Information for additional cases of misdiagnosed PM in our hospital was presented in Figure 1b and Tab S1.

### Performance of our PM-CAD system versus other established CNN models

We evaluated the diagnosis performance of 6 established CNN models as well as our PM-CAD system. Overall, in validation dataset, our PM-CAD system outperforms those 6 established CNN models (*i.e*., GoogLeNet ^[21]^, 3D-CNN ^[22]^, VGG ^[23]^, ResNet ^[24]^, DenseNet ^[25]^ and ResNeXt ^[26]^) (Tab 1, Fig S1 and Fig S2). Specifically, scores of AUC, F1, accuracy, positive predictive value (PPV) and negative predictive value (NPV) of PM-CAD system were 98.13%, 92.09%, 94.36%, 87.67%, and 98.36%, respectively (Tab 1). Then we compared those parameters with currently available well-established 6 CNN models, the 6 models also scored well, all of them the AUC above 95% and the diagnosis accuracy above 90% (Tab 1, Fig S1 and Fig S2). As we showed that performance of our PM-CAD is superior to 6 established CNN models. We then employed our PM-CAD system for the following investigation.

**Table 1.** Compare performance of PM-CAD system versus 6 established CNN models. We evaluate the performance of different models by using a series of metrics including the area under receiver operating characteristic curve (AUC) score (which is independent of probability threshold), F1-score, accuracy, sensitivity, positive predictive value (PPV), negative specificity and negative predictive value (NPV). Further information for each specific metric has been listed in Supplementary information for Materials and Methods, B.3. Experiment - Evaluation metric.

|  | AUC | F1-score | Accuracy | Positive Metrics |  | Negative Metrics |  |
| --- | --- | --- | --- | --- | --- | --- | --- |
|  |  |  |  | Sensitivity | PPV | Specificity | NPV |
| GoogLeNet | 0.9676 | 0.8857 (124/140) | 0.9179 (179/195) | 0.9394 (62/66) | 0.8378 (62/74) | 0.9070 (117/129) | 0.9669 (117/121) |
|  | [0.9390~0.9894] | [0.8210~0.9332] | [0.8702~0.9524] | [0.8520~0.9832] | [0.7339~0.9133] | [0.8431~0.9510] | [0.9175~0.9909] |
| 3D-CNN | 0.9686 | 0.8767 (128/146) | 0.9077 (177/195) | 0.9697 (64/66) | 0.8000 (64/80) | 0.8760 (113/129) | 0.9826 (113/115) |
|  | [0.9433~0.9886] | [0.8122~0.9253] | [0.8580~0.9444] | [0.8948~0.9963] | [0.6956~0.8811] | [0.8064~0.9274] | [0.9386~0.9979] |
| VGG | 0.9689 | 0.8841 (122/138) | 0.9179 (179/195) | 0.9242 (61/66) | 0.8472 (61/72) | 0.9147 (118/129) | 0.9593 (118/123) |
|  | [0.9449~0.9877] | [0.8186~0.9323] | [0.8702~0.9524] | [0.8320~0.9749] | [0.7431~0.9212] | [0.8526~0.9567] | [0.9077~0.9867] |
| ResNet | 0.9728 | 0.8936 (126/141) | 0.9231 (180/195) | 0.9545 (63/66) | 0.8400 (63/75) | 0.9070 (117/129) | 0.9750 (117/120) |
|  | [0.9508~0.9898] | [0.8306~0.9392] | [0.8763~0.9563] | [0.8729~0.9905] | [0.7372~0.9145] | [0.8431~0.9510] | [0.9287~0.9948] |
| DenseNet | 0.9733 | 0.8921 (124/139) | 0.9231 (180/195) | 0.9394 (62/66) | 0.8493 (62/73) | 0.9147 (118/129) | 0.9672 (118/122) |
|  | [0.9529~0.9888] | [0.8283~0.9383] | [0.8763~0.9563] | [0.8520~0.9832] | [0.7464~0.9223] | [0.8526~0.9567] | [0.9182~0.9910] |
| ResNeXt | 0.9740 | 0.9160 (120/131) | 0.9436 (184/195) | 0.9091 (60/66) | 0.9231 (60/65) | 0.9612 (124/129) | 0.9538 (124/130) |
|  | [0.9480~0.9935] | [0.8547~0.9573] | [0.9013~0.9715] | [0.8126~0.9659] | [0.8295~0.9745] | [0.9119~0.9873] | [0.9022~0.9829] |
| PM-CAD<br>(ours) | 0.9813 | 0.9209 (128/139) | 0.9436 (184/195) | 0.9697 (64/66) | 0.8767 (64/73) | 0.9302 (120/129) | 0.9836 (120/122) |
|  | [0.9625~0.9953] | [0.8628~0.9598] | [0.9013~0.9715] | [0.8948~0.9963] | [0.7788~0.9420] | [0.8717~0.9676] | [0.9420~0.9980] |

### Diagnosis performance of our PM-CAD system versus radiologists

An independent test A (100 participants: 50 PM patients and 50 controls) was used to compare our PM-CAD system with radiologists in our hospital. Six general radiologists were instructed to make a diagnosis for each MRI image. We evaluated the important parameters of diagnosis performance including F1 score, accuracy, sensitivity, PPV and NPV. We showed that the accuracy of our PM-CAD system is comparable to radiologist with more than 10 years of professional experience. The performance of our PM-CAD system is F1-score (93.88%), accuracy (94.00%), sensitivity (92.00%), PPV (95.83%), specificity (96.00%) and NPV is (92.31%) (Tab S2). In contrast, the performance of our best radiologist #6 is F1-score (94.95%), accuracy (95.00%), sensitivity (94.00%), PPV (95.92%), specificity (96.00%) and NPV is (94.12%) (Tab S2). The receiver operating characteristics (ROC) curves (showing both true-positive rate and false-positive rate for diagnosis performance) achieved on Test A are shown in Figure 2a. The AUC of PM-CAD system was 95.56%, and outperformed other five radiologists. At the same false-positive rate, the true positive rate of the PM-CAD system is higher than results of other 5 radiologists (Fig 2a). A false-positive result may cause unnecessary examination or excessive treatment. Moreover, a false-negative result may delay treatment and results in irreversible loss. To account for these issues, weighted error scoring ^[10]^ was incorporated during modelling and evaluation by radiologists. The PM-CAD system produces a weighted error of 10.00%, which is far below the average weighted error of 21.67% achieved by 6 radiologists (Fig 2b). The difference of negative or positive likelihood ratios ^[10]^ between our PM-CAD system and radiologists has achieved statistical significance (Fig 2c, d). Thus, our PM-CAD system demonstrates a better diagnostic accuracy than those of 5 radiologists in our hospital. The performance of PM-CAD system and radiologists are further evaluated by using the standard confusion matrices (Tab S3), with the number of true positives, false-positives, true negatives, and false-negatives being reported. We showed that PM-CAD system has obvious advantages in the diagnosis of PM than those of radiologists.

**Fig 2.**
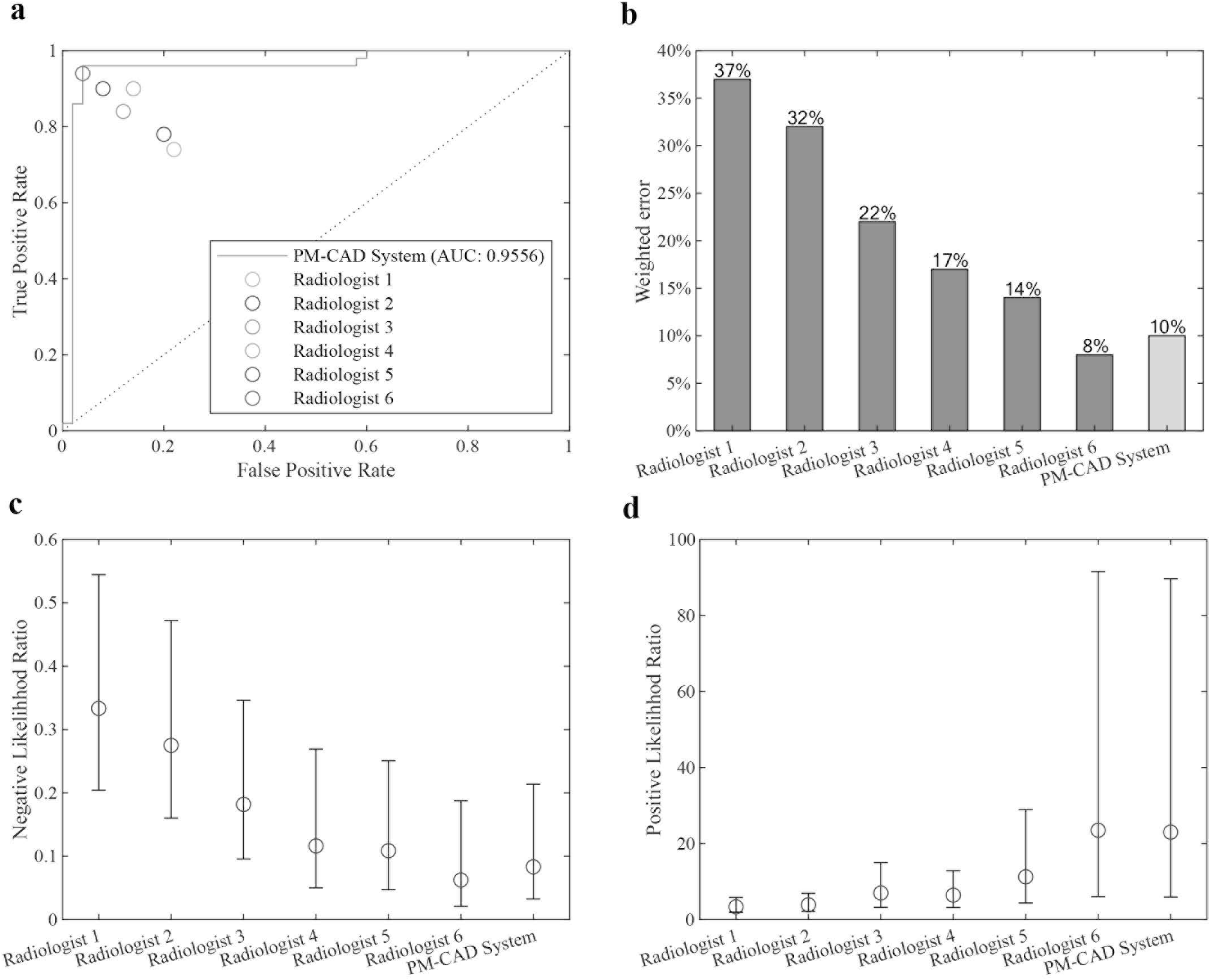
The PM-CAD system outperforms 5 radiologists in diagnosing PM. (a) ROC and AUC: ROC curve shows the true positive rates (sensitivity) with respect to different false-positive rates (1-specificity). The ROC results show that the PM-CAD system outperforms 5 radiologists. The area under ROC curve (AUC) of PM-CAD system is 95.56%. (b) Weighted error. A penalty weight of 2 is applied to false-negatives and a penalty weight of 1 is assigned to false-positives. The PM-CAD system produces a weighted error of 10%, whereas the radiologists produce a weighted error of 26.67%. (c & d) The negative likelihood ratio and the positive likelihood ratio: The negative likelihood ratio is defined as the false-negative rate over the true negative rate, so that a decreasing likelihood ratio < 1 indicated increasing probability the absence of PM. The positive likelihood ratio is defined as the true positive rate over the false-positive rate, so that an increasing likelihood ratio > 1 indicated increasing probability the diagnosis of PM. The confidence intervals show that the PM-CAD system demonstrates statistically better screening performance in terms of both negative likelihood ratio and positive likelihood ratio than radiologists. Radiologist 1 & 2: with < 5 years professional experience, Radiologist 3 & 4: with 5 - 10 years professional experience, Radiologist 5 & 6: with > 10 years professional experience. PM, pituitary microadenoma; receiver operating characteristics (ROC); the area under ROC curve (AUC).

### Further assessment for the accuracy of the PM-CAD system

We evaluated the accuracy of PM-CAD system in detecting positive cases and negative cases of PM. Timely diagnosis and treatment of positive cases can significantly improve disease prognosis. We sampled three double positive cases (both diagnosed by radiologists as well as PM-CAD system). Which undergoing surgical treatment, the double positive cases were confirmed by a subsequent pathological examination (Fig 3a and Tab S1). A false-negative result could lead to delay in treatment. We tested whether PM-CAD system could detect the false-negative cases of PM previously misdiagnosed by radiologist. We utilized the MRI dataset from 3 clinically misdiagnosed PM cases. Specifically, these 3 patients underwent surgical treatment (2 cases of Cushing’s disease and 1 case of TSH secreting adenoma). The presence of PM on those patients was confirmed by a subsequent pathological evaluation of the extracted tissue samples as well as other relevant clinical information (Fig 3b and Tab S1). To our delight, PM-CAD system was able to detect the presence of PM in all 3 previous misdiagnosed cases.

**Fig 3.**
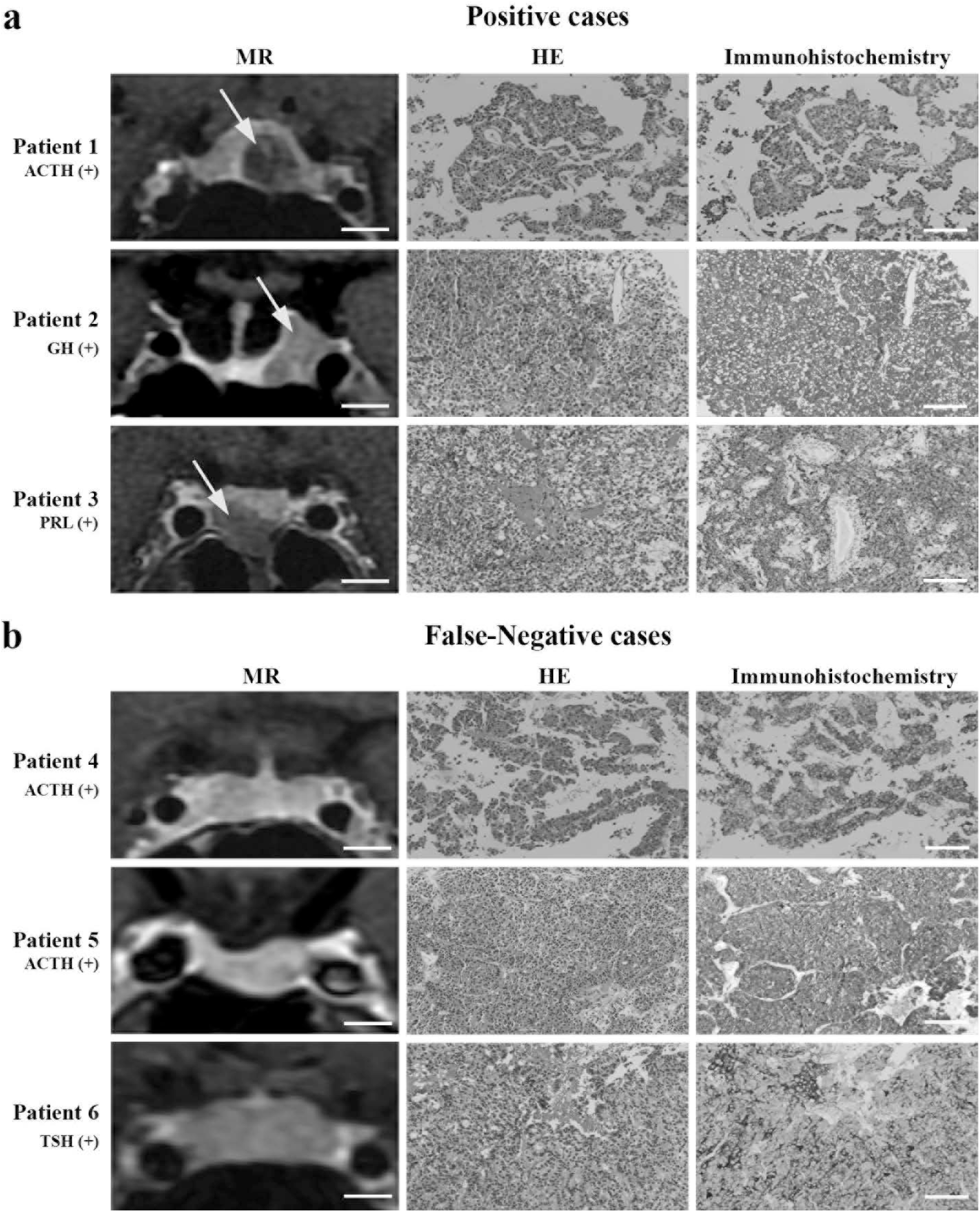
The MRI and pathologic images of double positive and false-negative cases. (a, b) 3 double positive and 3 false-negative cases, which were functional PM, as confirmed by subsequent pathological examination. (b) The PM-CAD system detects 3 false-negative cases previously misdiagnosed by radiologists. The comprehensive clinical data for these 3 misdiagnosed patients are listed in Supplementary Table 1. PM: pituitary microadenoma. MRI: magnetic resonance imaging. AI: Artificial intelligence. HE: hematoxylin and eosin. ACTH: adrenocorticotropic hormone. GH: growth hormone. TSH: thyroid stimulating hormone. PRL: prolactin. MR bar = 5mm. Pathology bar =100 μm. The yellow arrow indicates a pituitary microadenoma.

### Application of our PM-CAD system for PM diagnosis

We used both internal and external dataset to test the robust generalization performance of our PM-CAD system. The performance of our system was further tested in additional 150 patients from 3 different hospitals. Six general radiologists from each hospital were recruited (2 radiologists with < 5 years professional experience, 2 radiologists with 5 - 10 years professional experience while additional 2 radiologists with > 10 years professional experience). We observed that the accuracy of respective PM diagnosis was positively correlated to the radiologist’s professional experience as well as the time allocated for each image reading (Fig 4a, b). The diagnostic accuracy achieved by radiologists with professional experience >10 years was above 90%. Meanwhile, the diagnostic accuracy was higher than 88% when the MRI reading time for each patient was over 45 seconds. In contrast, the diagnosis accuracies achieved by PM-CAD system in internal hospital was 94 % and in external hospitals were 90 % and 88 %, respectively (Fig 4a). The results in the external datasets are very encouraging, we showed that the diagnosis performance of our PM-CAD system is comparable to radiologists with 5-10 years of professional experience and has negligible of time cost.

**Fig 4.**
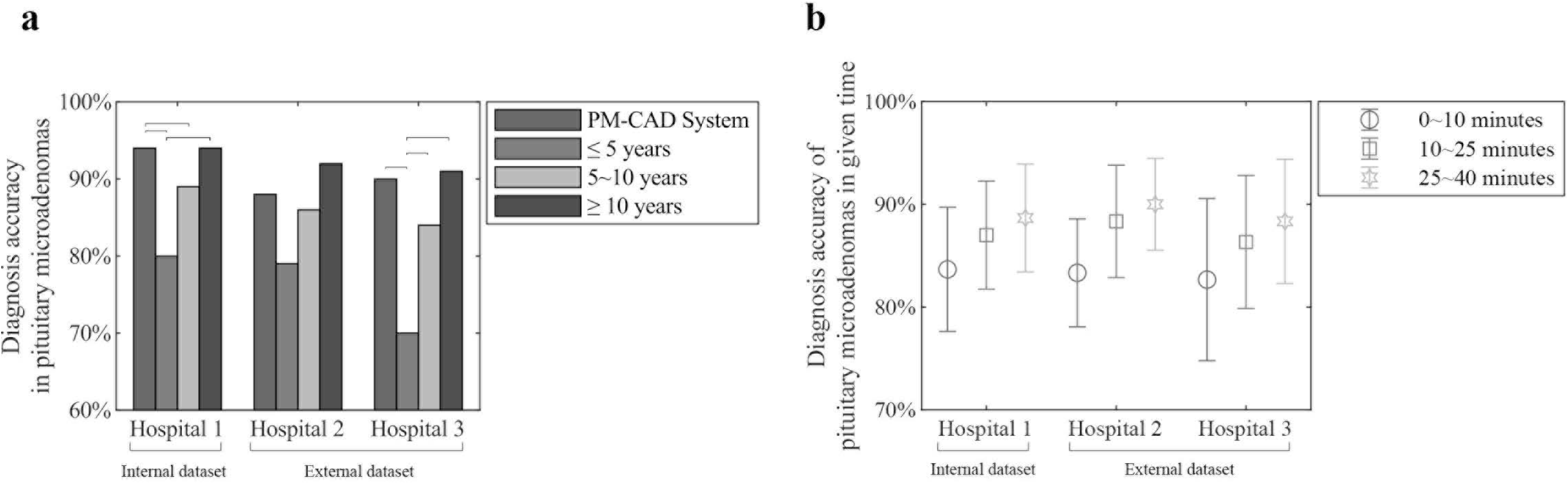
Application of the PM-CAD system in diagnosing PM. MR images are obtained from 3 hospitals. (a) The diagnosis accuracy rate is significantly increased in senior radiologist and the diagnosis accuracy of PM-CAD system is comparable to those radiologists >10 years of professional experience. (b) The diagnosis accuracy is significantly increased by radiologists with longer reading times allocated for each MRI scan. Bar means p < 0.05 for comparison of the two groups. PM: pituitary microadenoma. AI: Artificial intelligence. MR: magnetic resonance.

## DISCUSSION

Deep learning has been used to diagnose various diseases ^[15-17]^. However, existing CNN based pituitary diagnosis models mainly focus on pituitary macroadenoma ^[18-20]^. Our work provides the first evidence in applying CNN model for pituitary microadenoma diagnosis. We showed that our PM-CAD system outperforms 6 existing established CNNs-based models (*i.e*., GoogLeNet ^[21]^, 3D-CNN ^[22]^, VGG ^[23]^, ResNet ^[24]^, DenseNet ^[25]^ and ResNeXt ^[26]^) in diagnosis of PM, achieves satisfactory predictions on the validation dataset, with an accuracy of 94.36%, a sensitivity of 96.97%, and a specificity of 93.02%. Diagnosis of PM requires its fine-grained feature in MR images. However, the aggressive downsampling of most modern 2D CNN models harms the forward propagation of fine-grained feature ^[27]^. Furthermore, a large training dataset is required to train neural network parameters ^[28]^. To address the issues faced by fine-grained feature learning and overfitting, we introduced an improved backbone and an attention module (further information is listed in Supplementary information for Materials & Methods, section B– microadenoma diagnosis model) to our PM-CAD system. We showed that our PM-CAD system is more suitable for microadenoma diagnosis than 6 existing CNN models.

We also showed that the PM-CAD system outperformed 5 general radiologists in PM diagnosis. The weighted error achieved by PM-CAD was 10.00%, which was much lower that by radiologists (26.67%). The AUC achieved by PM-CAD system was 95.56% and was comparable to our radiologist with > 10 years of professional expertise (Fig 2a). By comparing the negative and positive likelihood ratio, we showed that the PM-CAD system achieved better diagnosis performance, with a higher positive diagnosis rate and a lower false-negative rate than radiologists.

To further confirm the diagnosis performance of PM-CAD system, three double positive cases (verified by PM-CAD system and radiologists) and three false-negative cases (misdiagnosed by radiologists) were selected. The diagnosis accuracy of PM-CAD system on false-negative cases was 100%, which was confirmed by a subsequent pathological examination. Three false-negative cases were hormone-producing microadenomas, with small size and irregular morphology which were difficult to detect by using MRI alone. The current MRI diagnostic sensitivity for hormone-producing primary and recurrent pituitary microadenomas was only 47% and 39% respectively ^[5]^. Our PM-CAD system demonstrates the excellent diagnosis performance in detecting those hormone-producing PM.

For this study, the MR images were collected from different MRI machines. It is important that our PM-CAD system achieves a robust generalization performance on images collected from different MRI machines. Images from 3 additional hospitals (1 internal and 2 external datasets) have been used to test the generalization diagnostic performance of PM-CAD system. The diagnostic accuracy achieved by the PM-CAD system in 3 hospitals were 94%, 90%, and 88%, respectively (Fig 4a). The internal test dataset and the training dataset have the same MR images sources, whereas the images in the external test dataset were generated with different MRI machines from different hospitals. Previous results suggest that CNN model performs better in the internal dataset than external dataset ^[29,30]^. Similar to those previous observations, we showed that PM-CAD system performs better on our internal test dataset than external dataset. Nevertheless, the diagnostic accuracy of PM-CAD system on external datasets was 90% and 88%, comparable to the diagnostic accuracy of radiologists with 5-10 years professional experience. We showed that our PM-CAD system could achieve robust generalization performance. The techniques presented in this study may potentially be applied to other hospitals in assisting for the diagnosis of PM.

MRI techniques for detection and depiction of pituitary adenomas have witnessed rapid evolution ranging from the non-contrast MRI scans to thin section contrast MRI scans ^[7,31]^. Modern MRI device has led to a rise in detection of PM as it allows evaluation of sella and perisellar lesions with high soft tissue contrast and excellent anatomical resolution ^[32,33]^. However, it is worth noting that accurate diagnosis of PM is still largely relied on the radiologist’s professional experience. As shown in Figure 4, the radiologist diagnostic accuracy for PM is proportionally enhanced with professional experience as well as allocated reading time for image. In this aspect, PM-CAD system has a distinct advantage in reading time and stability.

This work provides the first deep learning system for the diagnosis of PM. Our PM-CAD system provides a diagnostic accuracy comparable to experienced radiologists with marginal deployment costs. We acknowledge that there are several limitations for the current investigation. First, this is a retrospective study on multi-medical centers. Further validation with larger and prospective study is needed for clinical applications. Second, although a total of 1,228 participants have been used for training and testing of our deep learning system, the dataset is still insufficient. The robustness and accuracy of deep learning models can further be increased with more training data.

In conclusion, results from this investigation have highlighted the potential applications of deep learning on the diagnosis of patients with PM. With the rapid development of computing power, deep learning algorithms can surpass gold diagnosis standard. Machine learning for the diagnosis of PM will serve as an important component in improving patient care and outcomes.

## MATERIALS AND METHODS

### Ethical Approval

This study is approved by the research ethics committee of the Institute of Basic Research in Clinical Medicine, The Third Affiliated Hospital of Sun Yat-Sen University ([2020]02-089-01). This research is registered at the Chinese Clinical Trials Registry (http://www.chictr.org.cn/index.aspx) with the number ChiCTR2000032762.

### Data collection and pre-procession of MRI data

A total of 1,228 participants (543 PM patients and 685 controls) with pituitary MR images (28,839 images) (acquired by GE or Philips machines) were selected from the retrospective cohorts of patients from 3 hospitals from January 2014 to December 2019. All these MR images were randomly assigned regardless of the type of MRI machine or manufacturer. The internal dataset was collected from The Third Affiliated Hospital of Sun Yat-Sen University (*i.e*., hospital 1) and was mainly used for training, validation and test A and B. The external dataset collected from The Sun Yat-Sen Memorial Hospital of Sun Yat-Sen University (*i.e*., hospital 2) and The Second Affiliated Hospital of Harbin Medical University (*i.e*., hospital 3) was used for Test C. Workflow diagram for the overall experimental design was in Fig 5. MRI was performed as a part of patients’ routine clinical care. There were no exclusion criteria based on age, gender or ethnicity. Local electronic medical record databases for the newly diagnoses PM (PM group) and normal pituitary (control group) were included. The coronal dynamic enhancement T1 -weighted imaging (T1WI) sequence of MRI (DICOM) was downloaded with a standard image format according to the manufacturer’s software and instructions. All pituitary images were read by 2 neuroradiologistes and 1 neurosurgeon who had > 10 years of professional experience, and the diagnosis agreed by all 3 doctors were proceed for further investigation in this study. These data were randomly divided into training, validation or test sets for further analysis. In the training and validation datasets, all images present PM were selected by 4 general radiologists (> 5 years of professional experience) and reviewed by 2 neuroradiologistes. All images of coronal dynamic enhancement T1WI sequence were used for test A, B and C datasets without additional human intervention. MRI was performed with a 1.5 or 3.0 T MRI unit (GE, Philips company) in the head-first supine position, 380 ms/12.5 ms (repetition time /echo time) and 1 or 3 mm-thick section. We excluded the MR images without complete pituitary scan or with too many MRI artifacts. 6 endocrinology fellows were involved in collecting patient clinical information, and the dataset were reviewed by 2 endocrinologists. Among PM patients, there were 391 cases of non-functional pituitary microadenomas and 152 cases of functional pituitary microadenomas (Fig S3). The MR image and relevant clinical information was retrieved from the digital database of respective hospitals.

**Fig 5.**
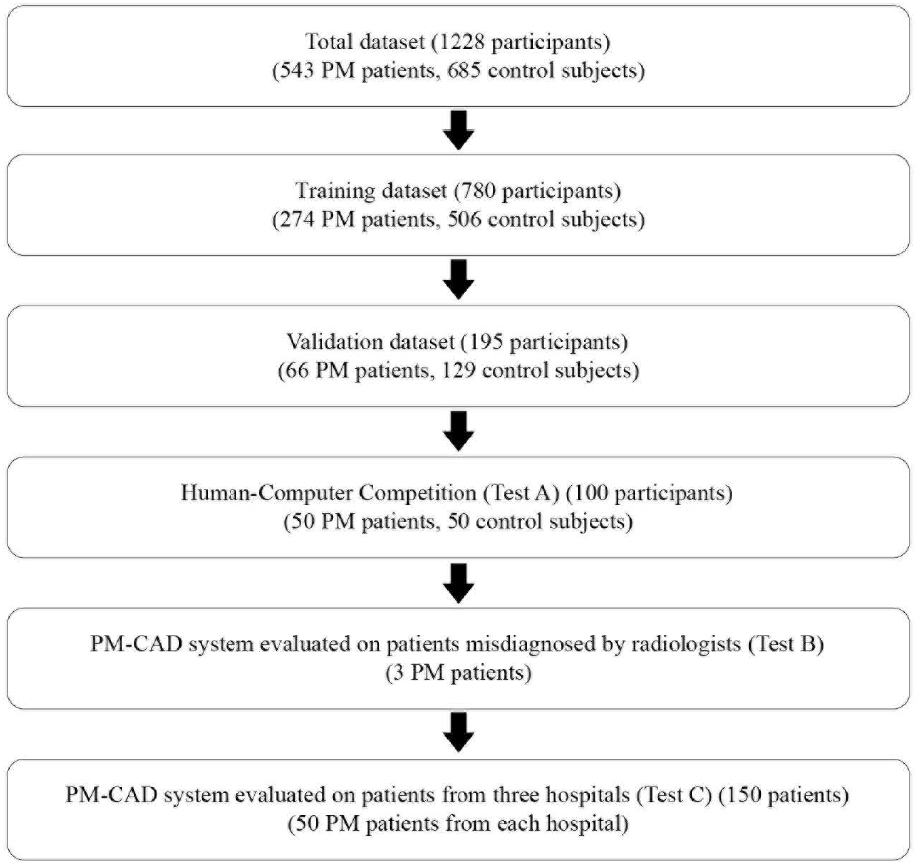
Workflow diagram for the overall experimental design. Pituitary MRI was performed as a part of patients’ routine clinical care. There were no exclusion criteria based on age, gender, or race. PM: pituitary microadenoma. MRI: magnetic resonance imaging.

### Classification of the dataset

All participants were split into 5 parts (the training dataset, the validation dataset, the test A, B and C dataset.). Workflow diagram for the overall experimental design was illustrated in Fig 5. The training dataset was used to train the deep learning models. It consists of 780 participants, including 274 PM patients and 506 normal controls, from the Third Affiliated Hospital of Sun Yat-Sen University. The validation dataset was used to tune the hyper-parameters (e.g., learning rate, number of training epochs) of the models and select the best model. It consists of 195 participants from the same hospital as the training dataset, including 66 PM patients and 129 normal controls. The test dataset was divided into three parts, namely Test A, B and C for the diagnosis performance comparison between radiologists and PM-CAD system. Test A consists of 100 participants (50 PM patients and 50 control subjests) from hospital 1 and has been used to comparing the PM diagnosis performance of PM-CAD system and general radiologists. There is no overlap among images in the training and validation datasets. Six radiologists were recruited for this study. Radiologists 1 and 2 has professional experience < 5 years, Radiologists 3 and 4 has professional experience for 5 to 10 years, Radiologist 5 and 6 has professional experience > 10 years. Each radiologist read 100 participants MR images independently in 50 minutes (about 30 second for each MRI). In test B, we tested the diagnosis performances of our PM-CAD system on 3 misdiagnosed cases of PM. Test C has been used to evaluate the generalization ability and stability of the PM-CAD system on MRI scan from three hospitals (hospital 1 was used to form the internal dataset, hospital 2 and 3 were used to form the external dataset) and each hospital provided 50 cases of PM images. Six general radiologists were also recruited to read MR images, and the diagnostic accuracy of those radiologists were evaluated.

### Overview of our PM-CAD system

The pipeline of our PM-CAD system is shown in Figure 6. Specifically, each section of a MR image was first diagnosed by our CAD system. The MRI scan was then determined as normal if all sections of this MR images were diagnosed as non-microadenoma. In the pituitary detection model, the Faster R-CNN framework ^[34]^ was employed to locate pituitary region from MR images. The input MR image was processed by this model to generate classification and regression maps, which were further used to extract the pituitary bounding box in MR image.

**Fig 6.**
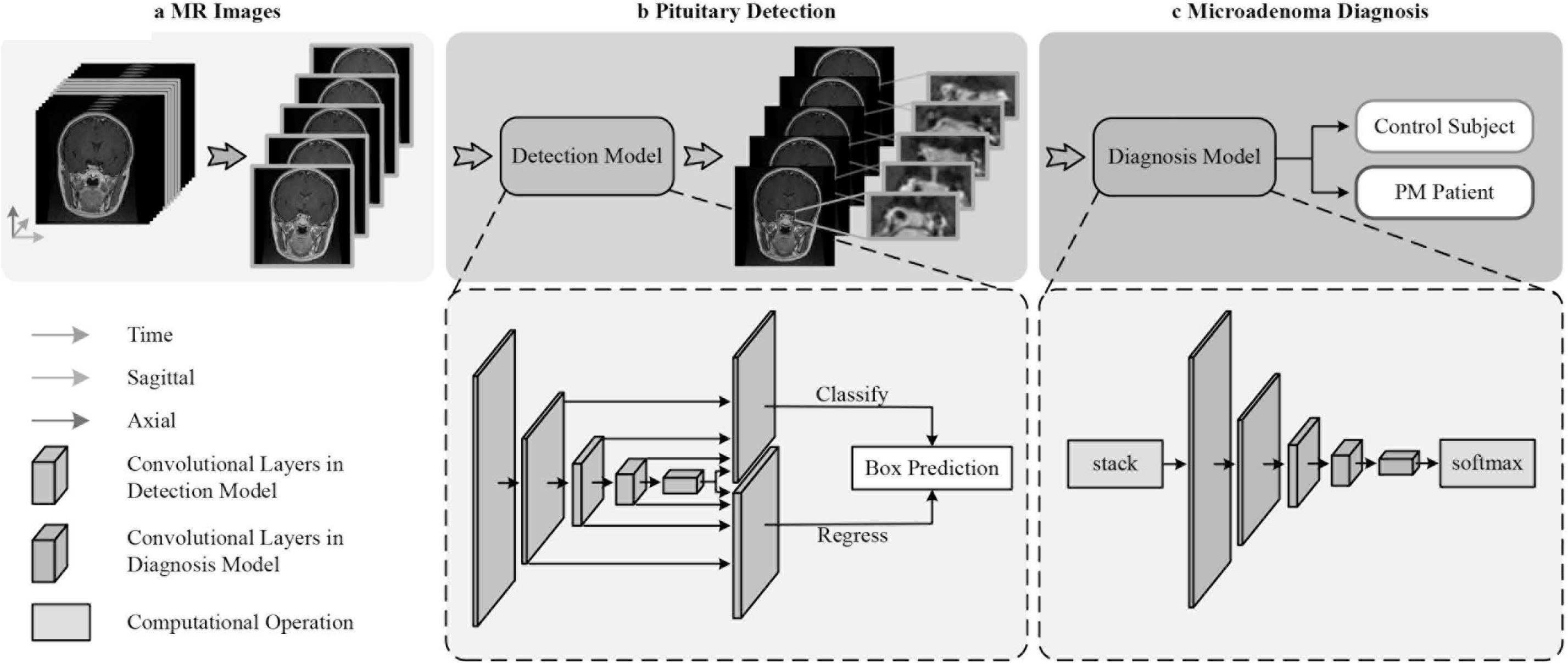
Overview of our PM-CAD system. (a) First the MR images are fed into our PM-CAD system for automatic diagnosis. The proposed PM-CAD system consists of two models: (b) the pituitary detection model localizes the pituitary region in cerebral MRI. The MR images are processed with multiple convolutional layers and two maps (classification map is used to predict the center and the regression map is used to refine the height and width of the rectangle box) are produced to predict a rectangle box enclosing the pituitary region. The pituitary rectangle region is cropped, stacked, and then fed into the PM diagnosis model. (c) it employs the proposed PM-Net model to extract features. A softmax layer is employed to transform the feature into the presence probability of PM. CAD: computer-aided diagnosis. MRI: magnetic resonance imaging. MR: magnetic resonance. PM: pituitary microadenoma.

**Fig 7.**
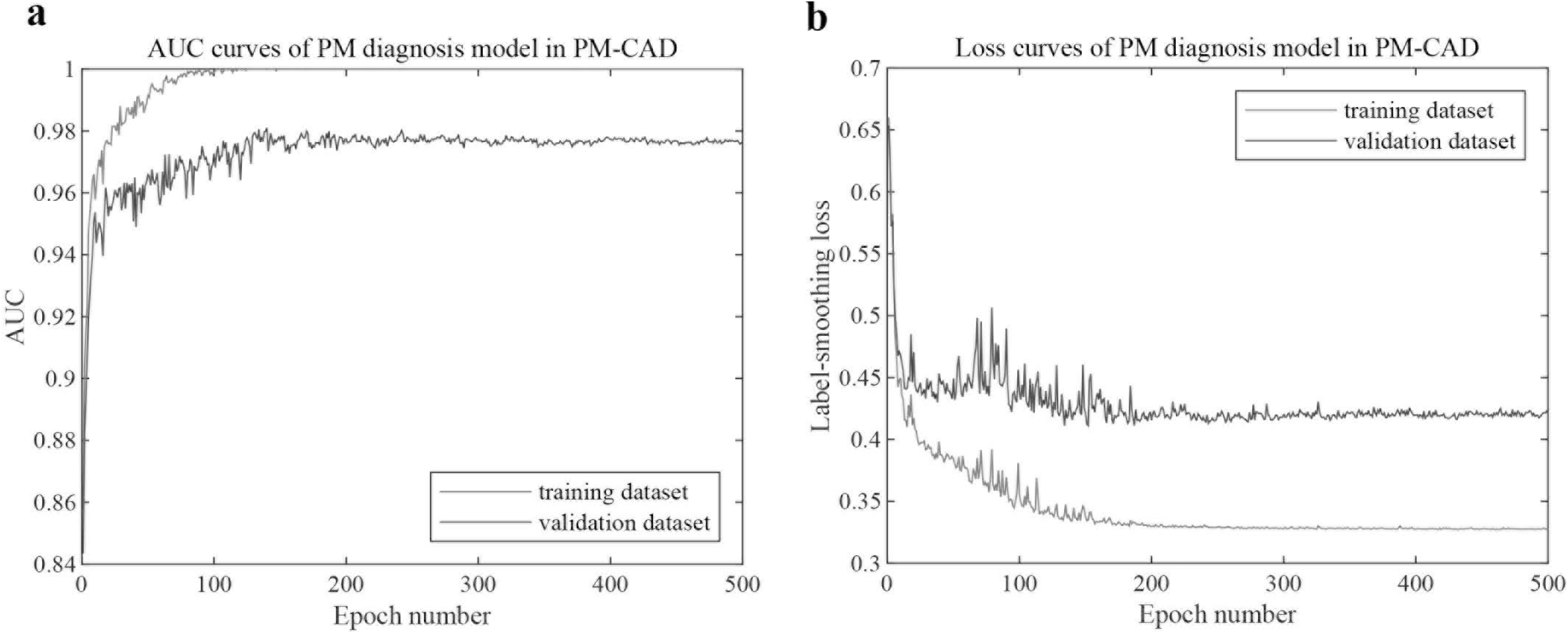
Performance of the PM-CAD system on the training and validation datasets. (a). Accuracy curves achieved by the PM-CAD system on the training and validation datasets. (b). Cross entropy loss curves achieved by the PM-CAD system on the training and validation datasets. We train the PM-CAD system for 500 epochs.

## Data Availability

All data have been presented in the manuscript and supplemental information.

## Supplemental information

### Supplementary information for Materials and Methods

#### A. Pituitary Detection Model

Although pituitary occupies a small region of MR images, radiologists can diagnose microadenoma based on the appearance of such a small region. On the other hand, directly feeding the whole MR images into a deep learning based diagnosis method imposes a huge challenge since non-pituitary regions will hinder microadenoma diagnosis significantly. Therefore, we employ a detection model to locate the pituitary region before diagnosis. The design of the detection model potentially benefits the consequence task in two aspects:

□ It enhances pituitary microadenoma (PM) feature by discarding irrelevant regions, and promotes the microadenoma diagnosis performance since the designed detection model can help the microadenoma diagnosis process focus on the pituitary region.
□ It reduces the overfitting problem of our microadenoma diagnosis model with a limited amount of data.

##### A.1. Architecture

Given several consecutive MR images at the same anatomical section of the brain from the coronal dynamic enhancement T1 -weighted imaging (T1WI) sequence of MRI scan, our pituitary detection model aims at locating the pituitary in each image. The pituitary detection model is build upon the Faster-RCNN ^[1]^ framework, and mainly consists of three parts: **ResNet-50 FPN** ^[2]^ extracts the multi-scale features from each image, **Region Proposals & Detection** produces the bounding box of pituitary from the multi-scale features, and **Post-processing** refines the detection results. (See Fig 1.)

**Figure 1.**
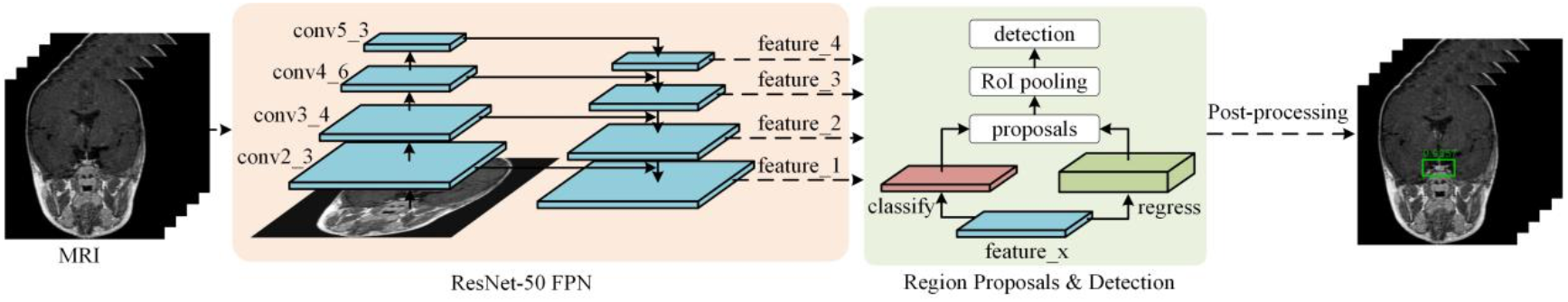
The pipeline of our pituitary detection model. The model consists of three prats: **ResNet-50 FPN** is used to extract the features in different scales, **Region Proposals & Detection** is used to generate the bounding boxes (which indicate pituitary regions), and **Post-processing** is used to refine the detection result.

###### ResNet-50 FPN

We use ResNet-50 FPN as our backbone to efficiently extract multi-scale features from each MR image. Specifically, ResNet-50 ^[3]^ is employed to process the input MR image, and four feature maps from conv2_3, conv3_4, conv4_6 and conv5_3 are further used as the input for the feature pyramid network (FPN). These four feature maps have 4×, 8×, 16×, 32× strided resolution compared to the input image.

FPN aggregates the above four feature maps, and produces four feature maps in different scales (i.e., feature_x where *x* ϵ{1,2,3,4}). The four feature maps are firstly processed by a 1×1 convolutional layer, to make them have the same number of channels. We denote the processed four feature maps as P_1, P_2, P_3 and P_4. The feature_4 is obtained by upsampling P4 with nearest interpolation. Feature_x (where *x* ϵ{2,3,4}) is obtained by upsampling the combination of the feature_(x-1) and the lateral feature map P_x. Therefore, four multi-scale features maps are used as the input for the following part.

###### Region Proposals & Detection

In this part, we process each feature map from ResNet-50 FPN independently, and several candidate bounding boxes indicating pituitary are produced. Specifically, given a feature map feature_x, two branch layers (each with one convolutional layer) are used to generate two feature maps. One branch is used to classify whether the anchor contains an object, and the another branch is used to regress the size (i.e., height, width) and position (i.e., x, y) from the default anchor. We obtain the bounding box proposals from these two feature maps using the non-maximum suppression (NMS) ^[1]^ algorithm to discard high-overlapping bounding boxes. Then, RoI pooling ^[1]^ is performed to aggregate the feature of feature_x within the bounding box proposals. The aggregated feature is further used to refine the classification and regression results, and produce several candidate bounding boxes of pituitary with confidence scores.

Each MR image has at most one pituitary. Therefore, from all candidate bounding boxes, we select the bounding box which has the highest confidence score. In order to reduce the false-positive rate of pituitary detection, we filter out the bounding box whose confidence score is smaller than a threshold *t* (*t* = 0.9 in our experiments).

###### Post-processing

Among all MR images, the above procedure might fail to locate the pituitary in some images. To handle this problem, we refine the pituitary detection result by post-processing. Specifically, we average all the positions and sizes of the detected bounding boxes, which can be considered as a mean bounding box. Because the position and size of pituitary over different MR images do not change a lot, we assign the missed detected MR image with the mean bounding box.

##### A.2. Experiment

###### Dataset

We collect 666 MRI scan (performed by GE, Philips machines) from the Third Affiliated Hospital of Sun Yat-Sen University. 2718 MR images with pituitaries from the coronal dynamic enhancement T1WI sequence of MRI are selected. A neuroradiologist annotates each image with a bounding box indicating the pituitary region. We randomly split all MRI scan into a pituitary detection training set with 532 scans (2167 images) and a pituitary detection validation set with 134 scans (551 images).

As the evaluation metrics, we use AP@0.50 and AP@0.75 from the COCO Detection Benchmark ^[4]^. AP@0.50 and AP@0.75 stand for the Average Precision (AP) with intersection of union (IOU) thresholds of 0.5 and 0.75, respectively.

###### Implementation

The pituitary detection model is implemented with PyTorch, and trained on a computer with an NVIDIA TITAN RTX GPU. We use the SGD optimizer with a momentum of 0.9 for training. The initial learning rate is set to 0.005. The warm-up strategy is adapted, which linearly increases the learning rate from 0.001 to the initial value over each iteration at the first epoch. Then, the learning rate decreases by 0.1 per 3 epochs. The weight decay is set to 0.0005. We resize the input images into 256×256 and normalize into (0,1) based on the window level (WL) and window width (WW) set by neuroradiologist as preprocessing. Other configurations (e.g., loss function, default anchor setting) are the same as ^[1]^. We train the pituitary detection model for 20 epochs with a batch size of 16.

###### Results

Table 1 shows the pituitary detection performance on MRI achieved by our method. On the training set, our method achieves 0.9884 and 0.9078 in terms of AP@0.50 and AP@0.75, respectively. The performance of pituitary detection on the validation set is close to that of the training set. That is, our model achieves a good generalization capability on unseen data. Moreover, our detection model can accurately locate the pituitary region. When 0.5 is used as the IOU threshold to determine the success of pituitary detection, our method achieves an average precision of 97.83% over different levels of recall.

**Table 1.** Pituitary Detection Results on MRI

|  | AP@0.50 | AP@0.75 |
| --- | --- | --- |
| Training set | 0.9884 | 0.9078 |
| Validation set | 0.9783 | 0.8824 |

Figure 2 shows the prediction results of our pituitary detection model and the ground-truth labels of several MR images on the validation set. The results demonstrate our method can produce accurate prediction bounding boxes with high overlaps with the ground-truth.

**Figure 2.**
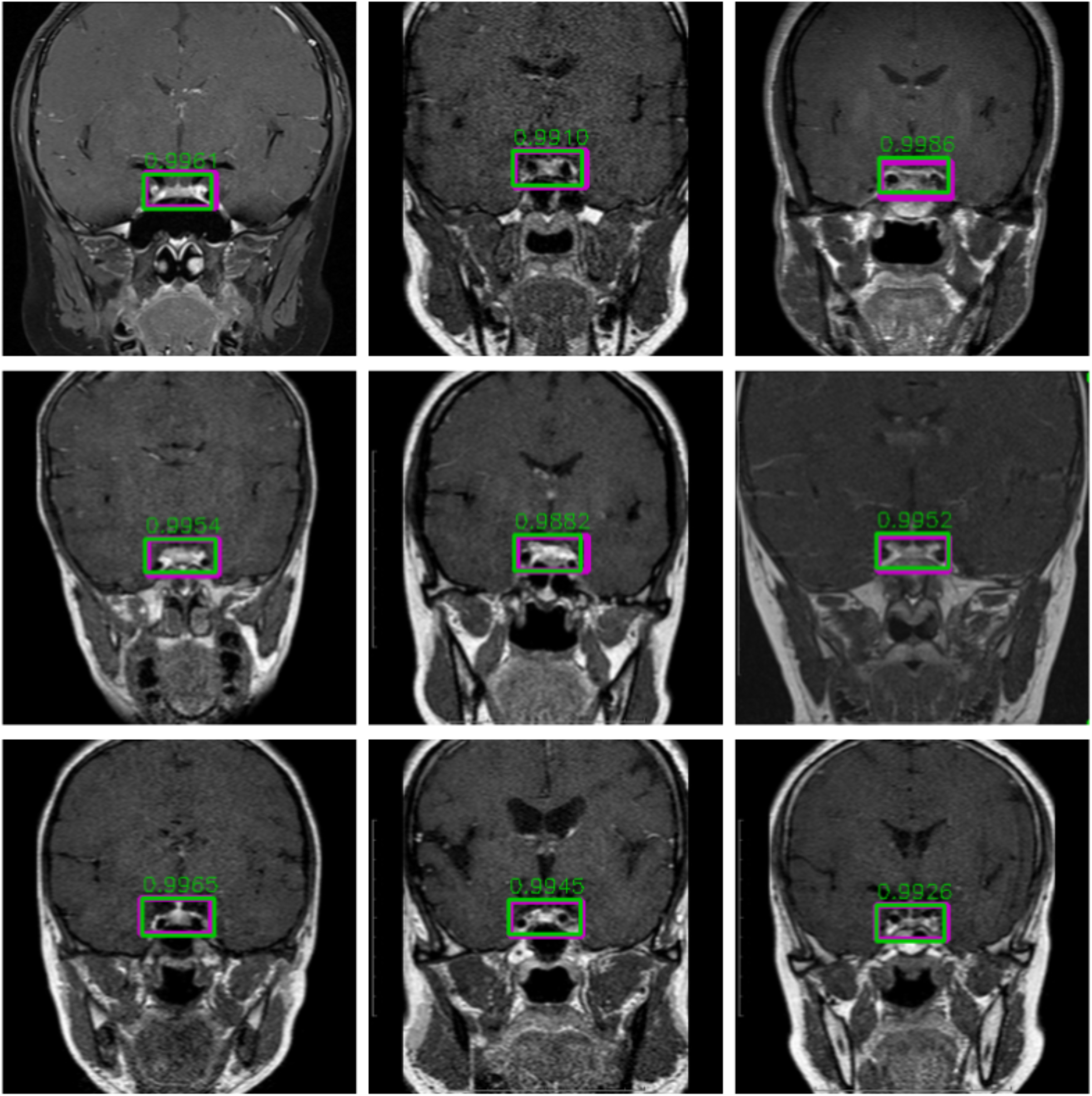
Some examples of pituitary detection on the validation set. Our method can produce accurate prediction bounding boxes (represented as green boxes) with high overlaps with the ground-truth (represented as purple boxes). Besides, the confidence scores produced by our pituitary detection model are also shown (i.e., the numbers near green boxes).

#### B. Pituitary Microadenoma Diagnosis Model

Automatic diagnosis of PM with deep learning is highly challenging due to three main reasons: (1) **Low inter-class variance**. Microadenoma occupies a very small part of the pituitary, the difference between microadenoma and normal tissues mainly lies in fine-grained textures. Actually, normal pituitaries and pituitaries with microadenoma are very similar in most parts, it is therefore very hard to identify microadenoma even by radiologist. Consequently, the proposed model should be powerful enough to distinguish these minor differences. (2) **High intra-class variance**. PM vary significantly in terms of location, size, and shape. Consequently, the proposed model should be sufficiently representative to adapt to these variations. (3) **Limited training data**. Deep learning techniques usually require a large amount of training data to avoid overfitting. However, the insufficiency of PM MRI scan poses new challenges to the design of architecture and training skills of neural networks.

To handle these challenges, we propose a novel CNN (namely, PM-CAD) for PM diagnosis. In PM-CAD, we modify the ResNet architecture to preserve fine-grained features during forward propagation. Furthermore, an attention module is used to further improve the discriminativeness of feature representation. To handle the overfitting problem, histogram matching normalization, intensity shift data augmentation and label-smoothing loss are used.

##### B.1. Architecture

**Figure 3.**
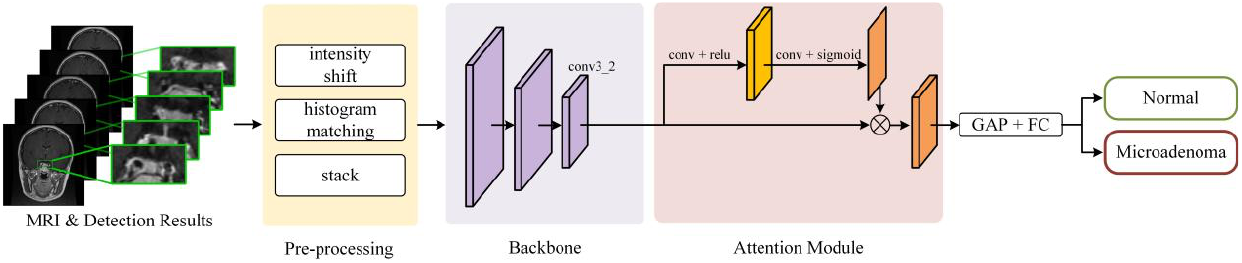
The pipeline of our microadenoma diagnosis model (PM-CAD). The model consists of three prats: **Pre-processing**, an improved **Backbone**, and an **Attention Module**. (conv: convolutional layer, relu: activation function, GAP: global average pooling, FC: fully connected layer.)

Given several consecutive MR images at the same anatomical position of the brain from the coronal dynamic enhancement T1WI sequence of MRI scan, our microadenoma diagnosis model aims at classifying them into normal or microadenoma. In particular, all MR images are processed using intensity shift as data augmentation and histogram matching normalization, then patches which only contain the pituitary are cropped from each MR image based on the detection results from the pituitary detection model. Then, all patches are resized and stacked as an image with multi-channels. Finally, our proposed CNN-based model processes these stacked pituitary patches and classifies them into normal or microadenoma.

In the following, we will first describe our CNN-based model with the improved **backbone** and an **attention module**, then highlight the intensity shift data augmentation and histogram matching normalization in **pre-processing**. Finally, we introduce the loss function used to train our CNN-based model.

###### Backbone

Most modern CNN models have two drawbacks when employing microadenoma diagnosis. (1) **Aggressive downsampling**. They use several max-pooling layers or strided convolutions to downsample the feature maps. For example, the ResNet series use a large kernel convolution with a stride of 2 followed by a max-pooling layer at the beginning, and 4 strided convolutional layers in the following. This strategy can efficiently reduce the computational cost and save the memory. However, fine-grained features which are important in PM classification might be lost in aggressive downsampling. (2) **Large amount of trainable parameters**. Modern CNN models tend to contain a large amount of trainable parameters. Therefore, a large training dataset is required to achieve a sufficient performance. However, the limited data of microadenoma MRI scan impose the over-fitting problem. A lightweight model with fewer parameters might be more suitable for microadenoma diagnosis.

Therefore, we extend ResNet-18 from two aspects. First, we replace the large kernel convolution with 3×3 convolution at the beginning of the network. Second, we remove the max-pooling layer at the beginning and the last stages of convolution to preserve fine-grained features. Therefore, *conv4_2* feature map is obtained with a 8× strided resolution. Then a global average pooling (GAP) and a multi-layer perceptron (MLP) with softmax are used to produce the probability of PM. Experimental results show that this simple modification of ResNet-18 can efficiently improve the performance of PM diagnosis.

###### Attention Module

The vector at each position of *conv4_2* feature map is considered as the feature of a small patch from the input. Since the number of normal patches is larger than that of microadenoma, the global average pooling (GAP) which directly performs on this feature map might make the microadenoma feature overwhelming from the normal feature. Therefore, we introduce an attention module to augment the microadenoma feature before GAP.

Given a feature map *f* of conv4_2, we employ a learnable convolution layer to produce a soft mask:

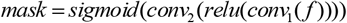

where *conv*_1_ and *conv*_2_ represent the 1×1 convolutional layers with 256 and 1 channels, respectively. This soft mask is further used to automatically select the prominent spatial areas of feature map *f* by pixel-wise multiplication as follows:

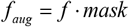

During training, this mask can learn to highlight the areas which have the discriminative features of microadenoma and suppress the irrelevant areas. Therefore, the augmented feature map contains the areas which is helpful for distinguishing the microadenoma and normal tissue.

The augmented feature map is further performed by GAP to aggregate spatial features. Then, the fully connected layers (FC) with softmax is used to produce the presence probability of microadenoma.

###### Pre-processing

Given several consecutive MR images with pituitary detection results, we first normalize them into (0,1) based on the window level (WL) and window width (WW). Then, data augmentation techniques such as Gaussian noise, translation, scaling, and the proposed intensity shift are performed during training. Furthermore, the proposed histogram matching normalization is used to align the histograms of all MR images. Finally, the patches only contain the pituitary are cropped from the MR images based on the detection result, and stacked as an image with mutli-channels. In the following, we will detail in our intensity shift data augmentation and histogram matching normalization.

###### Intensity shift data augmentation (IS)

To facilitate the domain knowledge from neuroradiologists, we normalize each MR image using WL and WW. This domain knowledge can help the network to focus on discriminative regions by assigning irrelevant pixels as background, and make the model converge faster. However, the normalization process decreases the generalization capacity on different hospitals. Different machines usually have different default values for WL and WW. Since WL and WW are adjusted by neuroradiologists based on their personal experience, the MR images normalized by different neuroradiologists from different machines in different hospitals have different distributions. Therefore, the model trained with the data from one hospital is difficult to generalize to other hospitals. To handle this limitation, we propose an intensity shift data augmentation approach to make the model insensitive to different WL and WW settings. In particular, we randomly shift the intensity of non-background pixels in MR images by adding a value within (0,0.1).

###### Histogram matching normalization (HM)

The MR images are acquired from the same anatomical section of brain. Therefore, the histograms between MR images are similar. In order to make the features between microadenoma and normal tissue more discriminative, we align the histograms using the histogram matching algorithm. Specifically, we select the histogram from one of the MR images as reference. Then, the histogram matching is performed to other MR image histograms to match the reference. In our experiments, we select the histogram from 3rd MR images among five consecutive MR images as the reference.

###### Loss

The cross-entropy loss is commonly used in classification. Specifically, the cross-entropy loss in binary classification can be formulated as:

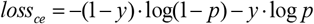

where *y* ∈{0,1} and*p* ∈(0,1)are the ground-truth label and the probability produced by the model. Usually, the probability is obtained by normalizing (e.g., softmax) the logits from the last layer of the model. The cross-entropy loss forces the logit to become an infinite value. That is, it pushes the distance of the learned feature between microadenoma and normal to be large, which potentially leads to over-fitting on the training data.

Motivated by ^[6]^, we use the label-smoothing loss as follows:

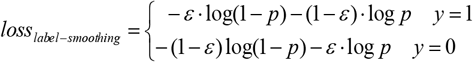

where *ε* is a small number and we set *ε* = 0.1 in our experiments. The label-smoothing loss can achieve the optimal performance when the logit is a finite value, this encourages the learned feature between microadenoma and normal more compact.

#### B.2. Application on MRI Scan

In this subsection, we will describe the approach for adapting our method to MRI scan.

Usually, a coronal dynamic enhancement T1WI sequence of MRI scan (DICOM) is used to diagnose PM. These images in a sequence can be divided into groups, and different groups focus on different anatomical sections of the pituitary. Each group contains several MR images, which are acquired at different enhancement times. 2 neuroradiologistes and 1 neurosurgeon examines all MR images of each pituitary section, and diagnoses whether microadenomas present in this section. The MRI scan is considered as normal if the microadenoma cannot be found in any pituitary sections.

Following the same idea, we adapt our PM-CAD to the MR images as to achieve end-to-end diagnosing without any human intervention. Specifically, the proposed method first inspects each section of the pituitary gland based on the learned feature, and then performs ‘OR’ operation based on the classification result of all sections. Similarly, the MR image is considered normal if and only if all sections of the pituitary are diagnosed as normal by our PM-CAD. Among MR images of each section, we select several consecutive images in the medium term of enhancement scan as the input for PM-CAD.

#### B.3. Experiment

##### Dataset

1228 participants with MRI scan were selected from retrospective cohorts of three hospitals from January 2014 to December 2019. The coronal dynamic enhancement T1WI sequence of MRI scan were used to diagnose PM in our experiments. All participants were split into 5 parts, that is:

**The Training Dataset**: It is used to train the deep learning models.

**The Validation Dataset**: It is used to tune the hyper-parameters (e.g., learning rate, number of training epochs) of the models and select the best model during training.

**Test A**: It is used to compare the performances between radiologists and our PM-CAD system in PM diagnosis.

**Test B**: It is used to test the diagnosis performances of our PM-CAD system on radilologists misdiagnosed cases.

**Test C**: It is used to evaluate the generalization ability of our proposed PM-CAD system on MRI scan from different hospitals.

##### Evaluation metric

To compare the performance of PM diagnosis between different models, we use the area under receiver operating characteristic curve (AUC) score (which is independent of probability threshold). In addition, F1-score, accuracy, sensitivity, specificity, positive predictive value (PPV), negative predictive value (NPV), error, positive likelihood ratio (PLR) and negative likelihood ratio (NLR) are used as metrics for performane evaluation.

The 95% confidence interval (CI) is also calculated for each metric. In particular, CI for AUC is calculated using bootstrap confidence intervals ^[10]^ (resample 50,000 times with replacement). CIs for F1-score, accuracy, sensitivity, specificity, PPV and NPV are calculated using the Clopper-Pearson interval ^[11]^ because it is common for calculating binomial confidence intervals. CIs for PLR and NLR are calculated using the “Log method” ^[7]^.

##### Implementation

We implement the microadenomas diagnosis model with PyTorch, and train our PM-CAD on NVIDIA TITAN RTX GPUs. We use the SGD optimizer with a momentum of 0.9. The initial learning rate is set to 0.02 and decreases by 0.99 per epoch. The weight decay is set to 0.0005. We train our PM-CAD for 500 epochs with a batch size of 16. We set the MLP in backbone to have two layers with 256 and 1 channels, respectively. The *ε* in our label-smoothing loss is set to 0.1. We fine-tune our PM-CAD model from the weights pre-trained on ImageNet ^[8]^.

The probability threshold of the model is selected based on the Youden Index ^[5]^ of the Receiver Operating Curve (ROC). Specifically, given the probabilities produced by the model and their corresponding ground-truth labels, we can first calculate the True Positive Rates (TPR) and the False Positive Rates (FPR) under different levels of threshold t. Then, the Youden Index at threshold t is defined as:

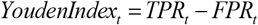

Finally, the threshold producing the largest Youden Index is considered as the optimal probability threshold 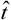:

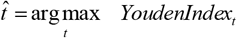

##### Comparison to existing methods

We compare our PM-CAD with several 2D and 3D baselines in PM diagnosis. In particular, five 2D CNNs-based models (i.e., GoogLeNet, VGG-16, ResNet-50, DenseNet-169 and ResNeXt-50 32×4d) are selected with the following considerations: these models achieve state-of-the-art performance on the largest image classification benchmark (i.e., ImageNet ^[8]^), and their superiority has been demonstrated in many other medical image analysis tasks. All these models are implemented in PyTorch. Since 5 MR images are stacked to form an image with multi-channels, the numbers of input channels and output categories are set to 5 and 2, respectively. To leverage the advantage of transfer learning, we also fine-tune these models from the weights pre-trained on ImageNet. We train these models with the same configuration as PM-CAD using the cross-entropy loss.

On the other hand, five consecutive MR images can be considered as a 3D volume. We employ a 3D CNN to diagnose PM from this volume. We choose 3D CNN from the Model Genesis ^[9]^, which is the first work serving as a primary source of transfer learning for 3D medical imaging applications. We fine-tune the 3D CNN model from the weights pre-trained on a large number of unlabeled medical images. We train it with the same configuration as PM-CAD using the cross-entropy loss.

We train the model using the training dataset, and select the best one which has the highest AUC score on the validation dataset during training. In manuscript, Table 1, Fig S1 and Fig S2 shows the results of our PM-CAD and existing models on the validation dataset.

##### Ablation Study

We provide the ablation study on the intensity shift data augmentation and the histogram matching normalization. We train our PM-CAD with or without intensity shift data augmentation and histogram matching normalization using training dataset, and evaluate it on Test C. Table 2 shows the accuracy has improved by 4% and 2% in hospital 3 and hospital 2, when equipped with HM and IS. And HM and IS do not improve the performance on the data from hospital 1. That is because, the training dataset and test data from hospital 1 have the same source which is the Third Affiliated Hospital of Sun Yat-Sen University, resulting in consistent normalization set by WL and WW. The result shows that the proposed HM and IS can eliminate the effect of inconsistent normalization from different hospitals. With HM and IS, our PM-CAD demonstrate a good generalization capacity, achieving above 88% accuracy of different hospitals.

**Table 2.** Accuracy of our PM-CAD (w/ or w/o HM, IS) on Test C (from 3 different hospitals).

|  | hospital 1 | hospital 2 | hospital 3 |
| --- | --- | --- | --- |
| w/o HM, IS | 47/50 | 44/50 | 42/50 |
| w/ HM, IS | 47/50 | 45/50 | 44/50 |

## Acknowledgments

This study was funded by National Key R&D Program of China (2017YFA0105803), the General Program of National Natural Science Foundation of China (U20A20185, 81770826, 61972435), the Key Area R&D Program of Guangdong Province (2019B020227003), the Natural Science Foundation of Guangdong Province (2019A1515011271), the Science and Technology Plan Projects of Guangzhou (202007040003), and the Science and Technology Innovation Committee of Shenzhen Municipality (JCYJ20190807152209394). We thank all the patients and investigators for their participation in this study. We thank Chunkui Shao and Jing Wang (Professor, Department of Pathology and radiology, The Third Affiliated Hospital of Sun Yat-Sen University) and The Magnetic Resonance Department of Second Affiliated Hospital of Harbin Medical University.

---

**Supplementary Table 1.**
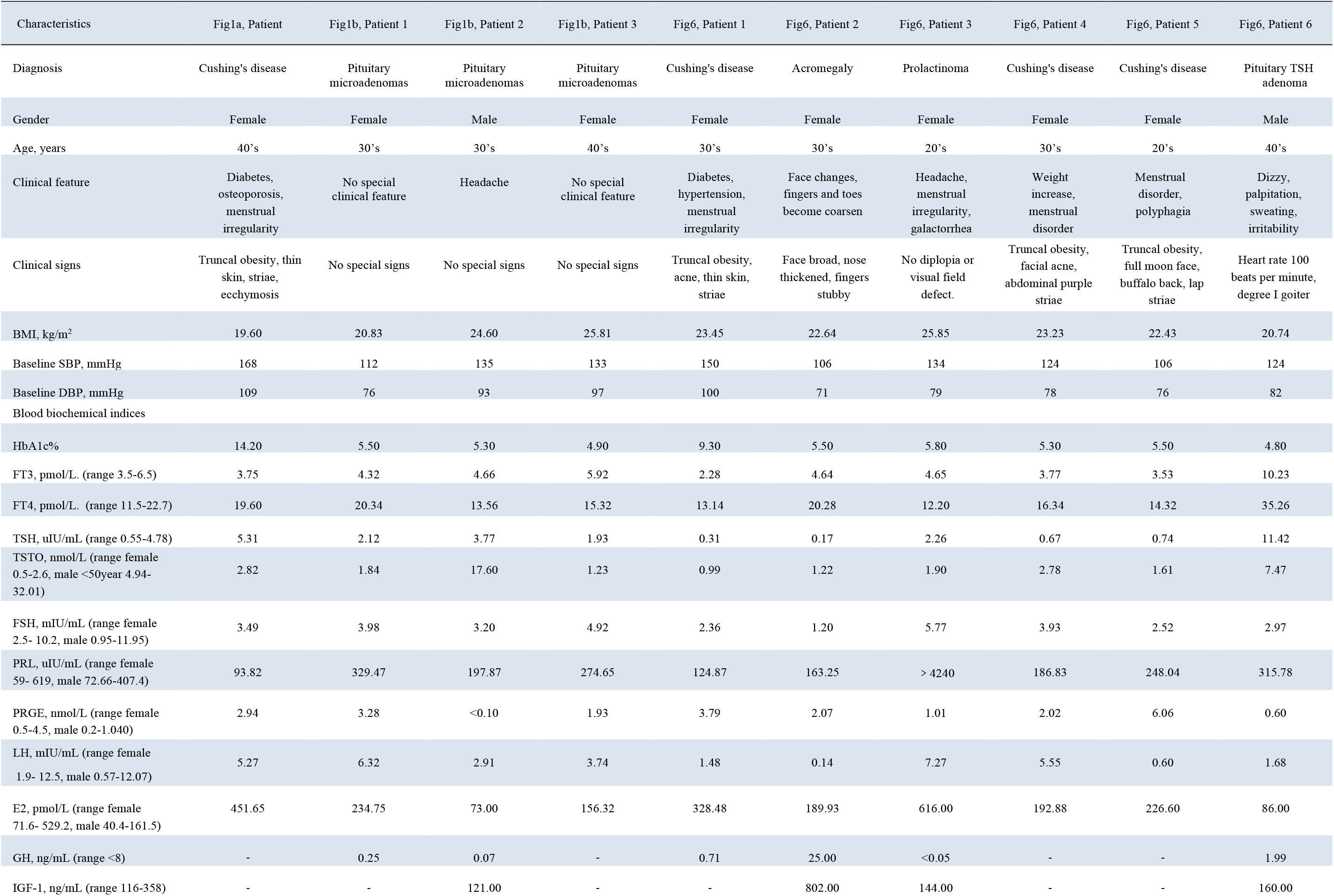
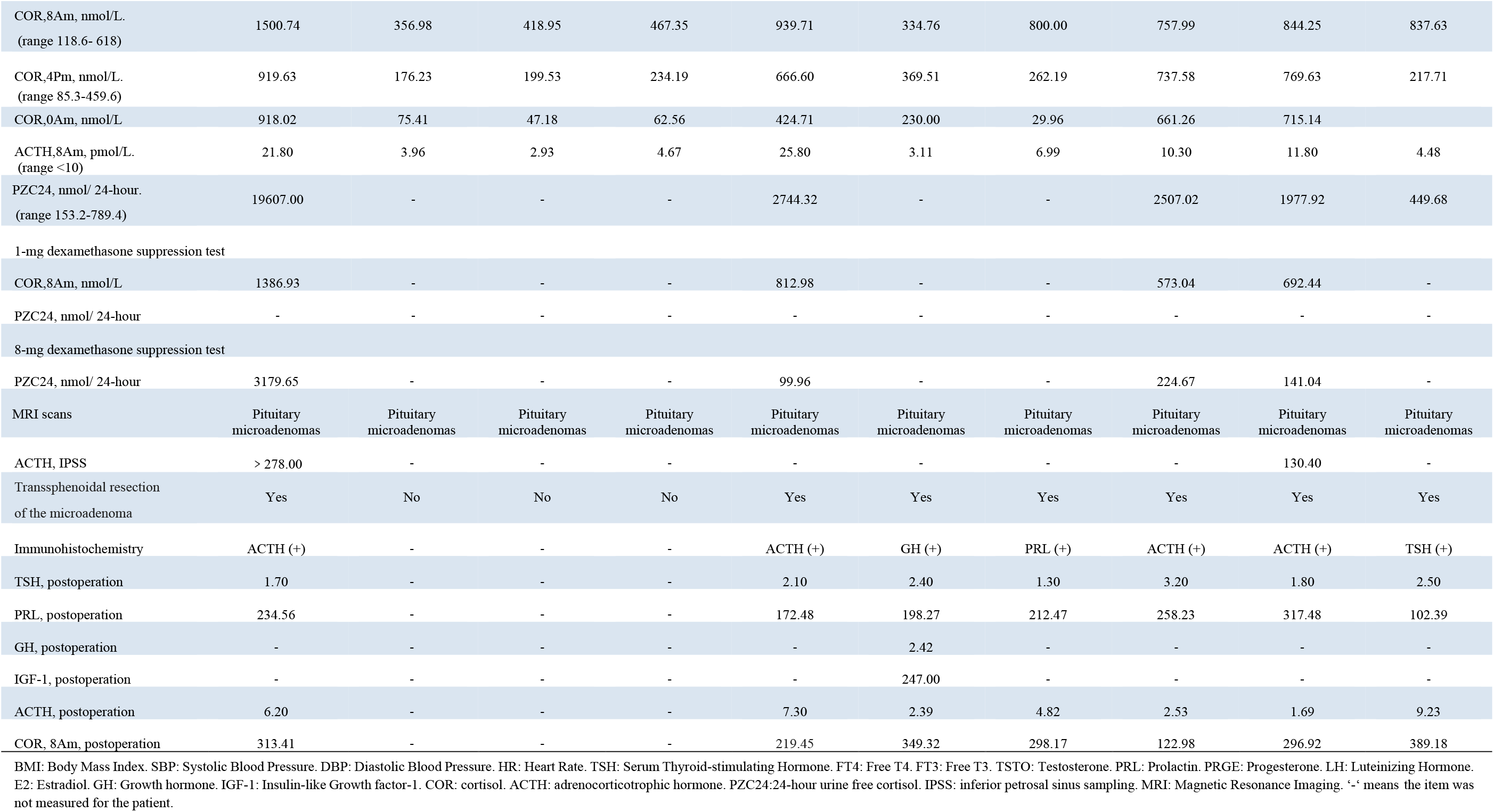
The patient clinical data in Fig 1 and Fig 3.

**Supplementary Table 2.** The diagnostic performance was comparable in our PM-CAD system versus radiologists (Test set A, n=100)

| Finding | PM-CAD | Radiologist 1 | Radiologist 2 | Radiologist 3 | Radiologist 4 | Radiologist 5 | Radiologist 6 |
| --- | --- | --- | --- | --- | --- | --- | --- |
| F1 score | 0.9388 (92/98) | 0.7551 (74/98) | 0.7879 (78/99) | 0.8571 (84/98) | 0.8824 (90/102) | 0.9091 (90/99) | 0.9495(94/99) |
|  | [0.8715~0.9772] | [0.6579~0.8364] | [0.6942~0.8636] | [0.7719~0.9196] | [0.8035~0.9377] | [0.8344~0.9576] | [0.8861~0.9834] |
| Accuracy | 0.9400 (94/100) | 0.7600 (76/100) | 0.7900 (79/100) | 0.8600 (86/100) | 0.8800 (88/100) | 0.9100 (91/100) | 0.9500(95/100) |
|  | [0.8740~0.9777] | [0.6643~0.8398] | [0.6971~0.8651] | [0.7763~0.9213] | [0.7998~0.9364] | [0.8360~0.9580] | [0.8872~0.9836] |
| Sensitivity | 0.9200 (46/50) | 0.7400 (37/50) | 0.7800 (39/50) | 0.8400 (42/50) | 0.9000 (45/50) | 0.9000 (45/50) | 0.9400(47/50) |
|  | [0.8077~0.9778] | [0.5965~0.8537] | [0.6400~0.8847] | [0.7089~0.9283] | [0.7819~0.9667] | [0.7819~0.9667] | [0.8345~0.9874] |
| PPV | 0.9583 (46/48) | 0.7708 (37/48) | 0.7959 (39/49) | 0.8750 (42/48) | 0.8654 (45/52) | 0.9184 (45/49) | 0.9592(47/49) |
|  | [0.8575~0.9949] | [0.6269~0.8797] | [0.6566~0.8976] | [0.7475~0.9527] | [0.7421~0.9441] | [0.8040~0.9773] | [0.8602~0.9950] |
| Specificity | 0.9600 (48/50) | 0.7800 (39/50) | 0.8000 (40/50) | 0.8800 (44/50) | 0.8600 (43/50) | 0.9200 (46/50) | 0.9600(48/50) |
|  | [0.8629~0.9951] | [0.6404~0.8847] | [0.6628~0.8997] | [0.7569~0.9547] | [0.7326~0.9418] | [0.8077~0.9778] | [0.8629~0.9951] |
| NPV | 0.9231 (48/52) | 0.7500 (39/52) | 0.7843 (40/51) | 0.8462 (44/52) | 0.8958 (43/48) | 0.9020 (46/51) | 0.9412(48/51) |
|  | [0.8146~0.9786] | [0.6105~0.8597] | [0.6468~0.8871] | [0.7192~0.9312] | [0.7734~0.9653] | [0.7859~0.9674] | [0.8376~0.9877] |

Note.—Unless otherwise specified, data are shown in percentages, with numbers of images in parentheses and 95% confidence intervals in brackets. F1 score: the harmonic mean of PPV and sensitivity. NPV: negative predictive value. PPV: positive predictive value. Radiologist 1 & 2: < 5 years professional experience; Radiologist 3 & 4: 5 - 10 years professional experience; Radiologist 5 & 6: > 10 years professional experience.

**Supplementary Table 3.** Confusion Matrices achieved by the PM-CAD system and radiologists on Test Set A (n=100)

| Prediction | PM-CAD (Truth) |  | Radiologist 1 (Truth) |  | Radiologist 2 (Truth) |  | Radiologist 3 (Truth) |  | Radiologist 4 (Truth) |  | Radiologist 5 (Truth) |  | Radiologist 6 (Truth) |  |
| --- | --- | --- | --- | --- | --- | --- | --- | --- | --- | --- | --- | --- | --- | --- |
|  | Adenoma | Normal | Adenoma | Normal | Adenoma | Normal | Adenoma | Normal | Adenoma | Normal | Adenoma | Normal | Adenoma | Normal |
| Adenoma | 46 (a) | 2 (b) | 37 | 11 | 39 | 10 | 42 | 6 | 45 | 7 | 45 | 4 | 47 | 2 |
| Normal | 4 (c) | 48 (d) | 13 | 39 | 11 | 40 | 8 | 44 | 5 | 43 | 5 | 46 | 3 | 48 |

Note.—Data are numbers of images. a: true-positive, b: false-positive, c: false-negative, d: true-negative.

## Legend for Supplementary figures

**Supplementary Fig 1.**
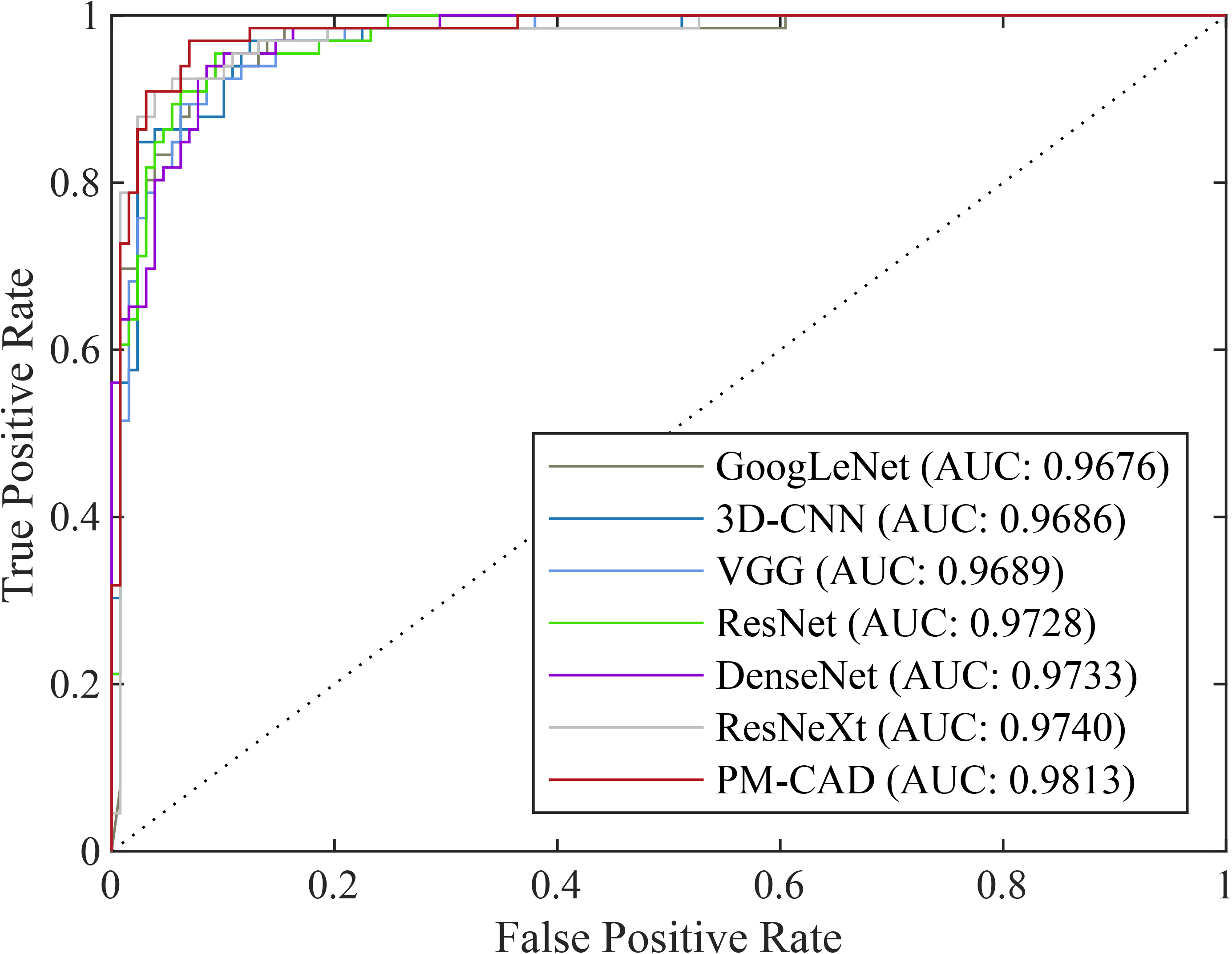
The ROC curves of different PM diagnosis models. Receiver operating characteristics (ROC) curves plot the true positive rate (sensitivity) versus the false positive rate (1 - specificity). The highest area under ROC curve (AUC) value is 0.9813 for PM-CAD. The AUC values for other 6 CNN models are also shown. CNN: convolutional neural network.

**Supplementary Fig 2.**
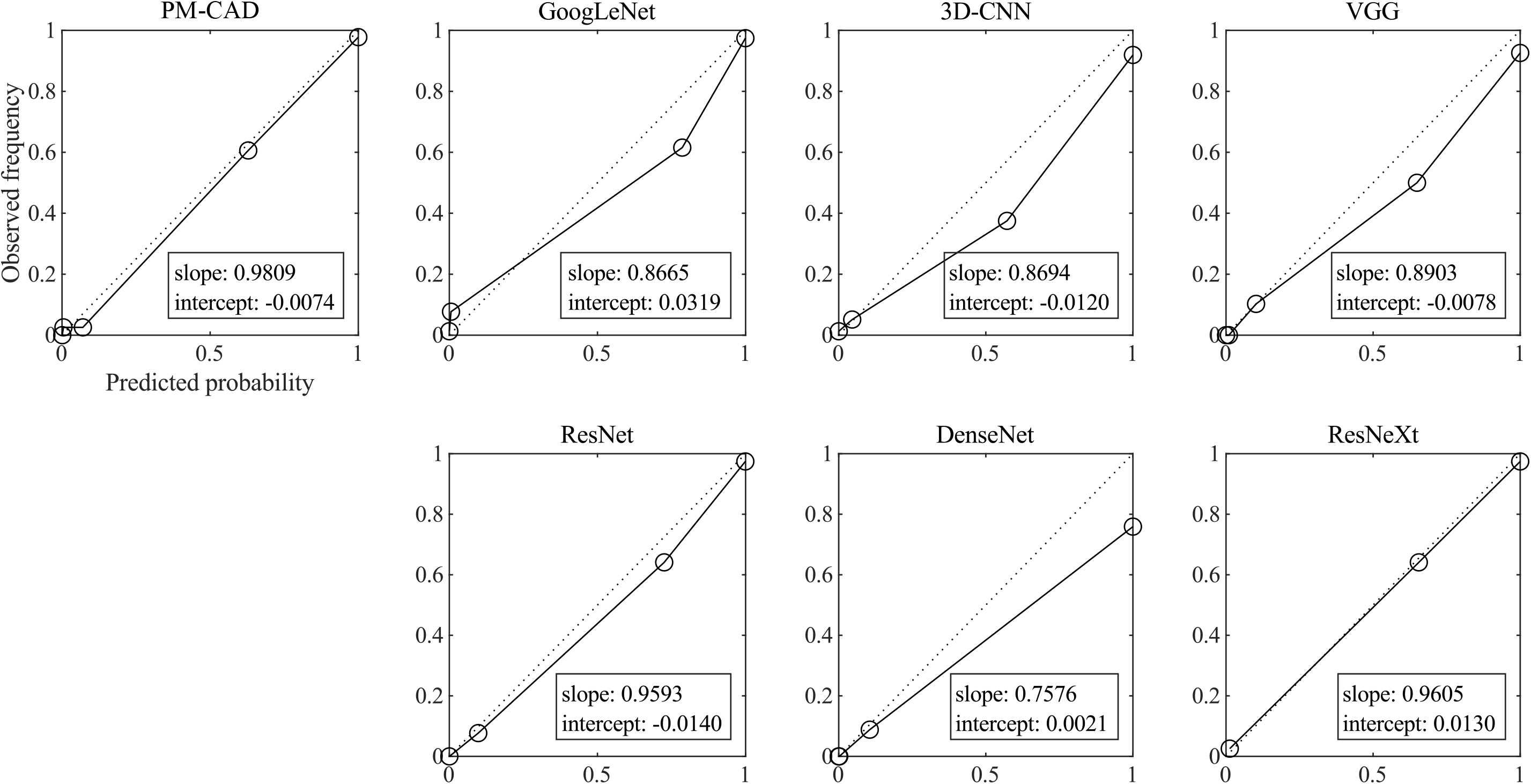
Calibration curves of different PM diagnosis models. Calibration curves for predicted versus observed risk in the overall validation cohort, PM-CAD shows optimal diagnostic performance.

**Supplementary Fig 3.**
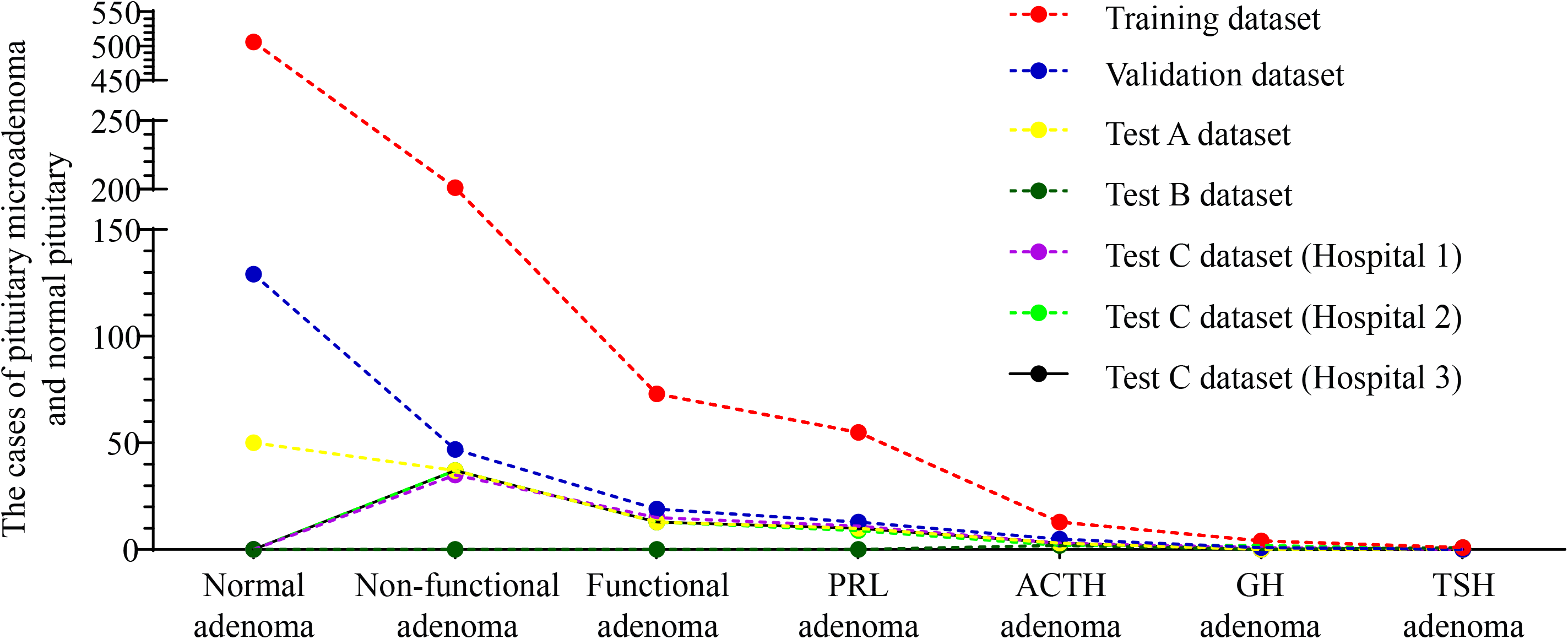
Data distribution diagram. The distribution of functional and nonfunctional pituitary microadenomas in the training, validation and test dataset.

